# Low-frequency repetitive transcranial magnetic stimulation in patients with motor deficits after brain tumor resection: a randomised, double-blind, sham-controlled trial

**DOI:** 10.1101/2023.03.14.23287254

**Authors:** Melina Engelhardt, Heike Schneider, Jan Reuther, Ulrike Grittner, Peter Vajkoczy, Thomas Picht, Tizian Rosenstock

## Abstract

**Background:** Surgical resection of motor eloquent tumors poses the risk of causing postoperative motor deficits which leads to reduced quality of life in these patients. Currently, rehabilitative procedures are limited with physical therapy being the main treatment option.

**Objective:** The present study investigated the efficacy of repetitive navigated transcranial magnetic stimulation (rTMS) for treatment of motor deficits after supratentorial tumor resection.

**Methods:** This randomised, double-blind, sham-controlled trial recruited patients with a worsening of upper extremity motor function after tumor resection. They were randomly assigned to receive rTMS treatment (1Hz, 110% RMT, 15 minutes, 7 days) or sham stimulation to the motor cortex contralateral to the injury followed by physical therapy. Motor and neurological function as well as quality of life were assessed directly after the intervention, one month and three months postoperatively.

**Results:** Thirty patients were recruited for this study. There was no significant difference between both groups in the primary outcome, the Fugl Meyer score three months postoperatively (Group difference [95%-CI]: 5.05 [-16.0; 26.1]; p=0.631). Patients in the rTMS group presented with better hand motor function (BMRC scores) one month postoperatively. Additionally, a subgroup of patients with motor eloquent ischemia showed lower NIHSS scores at all timepoints.

**Conclusion:** Low-frequency rTMS facilitated the recovery process stimulated hand muscles, but with limited generalization to other functional deficits. Long-term motor deficits were not impacted by rTMS. Due to the reduced life expectancy in this patient group a shortened recovery duration of functional deficits can still be of high clinical significance.

## Introduction

The resection of a tumor in vicinity to motor eloquent cortical or subcortical areas bears the risk to induce postoperative motor deficits. A worsening of motor function after surgery is observed in approximately one fourth of these patients. A full recovery is not always possible in the following months [1,2]. These functional deficits contribute to an overall impaired neurological status and are directly associated with a decreased life expectancy and a reduced quality of life [3]. Hence, improving neurorehabilitation of new deficits offers great potential to increase quality of life in these patients.

Healthy motor function critically depends on the communication between the motor cortices of both hemispheres. Specifically, transcallosal motor signals are used to control bimanual movements as well as to inhibit mirror movements during unimanual movements [4]. In patients with unihemispheric lesions, this communication can be altered which has been associated with motor deficits and a reduced motor recovery. Research in stroke patients shows an increased excitability of the unaffected hemisphere together with an increased transcallosal inhibition towards the affected motor areas [5,6]. These processes limit the recovery of the affected hemisphere and might have a negative impact on motor deficits. Consequently, methods to normalize transcallosal communication have the potential to improve motor symptoms.

Low-frequency repetitive transcranial magnetic stimulation (rTMS) can downregulate activity of stimulated brain areas noninvasively by applying a series of magnetic pulses to the scalp [7]. Application of low-frequency rTMS is further associated with an increase in activity of the contralateral motor cortex and has been successfully used to facilitate motor recovery in stroke patients [8]. Specifically, the combination of individualized physical therapy and rTMS seems to lead to favorable outcomes.

In brain tumor patients, early physical training is associated with a significant improvement of neurological function [9]. In a subgroup of tumor patients with subcortical ischemia within the corticospinal tract and preserved motor evoked potentials, this effect was further enhanced with rTMS as an additive treatment [10]. However, the inclusion criteria of this study limit the potential target population of brain tumor patients that could benefit from the intervention significantly.

Therefore, we aimed to investigate generalizability off the previously stated effects of combined low-frequency rTMS and physical therapy on recovery of postoperative motor deficits in a larger group of brain tumor patients. Further, we included health-related quality of life as important patient-related outcome.

## Materials and Methods

### Study design

In this randomised, double-blind, sham-controlled, monocentric parallel-group two-arm phase 2 trial (DRKS00010043) brain tumor patients were recruited from the neurosurgical ward at Charité – Universitätsmedizin Berlin, Germany between 2016 and 2021. The trial was approved by the local ethics committee at Charité (EA4/132/15) and conducted in accordance with the Declaration of Helsinki [11].

### Patients

Patients with a supratentorial brain tumor were eligible for study participation if they presented with a worsening of upper extremity motor function after surgical resection of the tumor as determined based on preoperative patient records. Eligible patients were either referred to the trial physician by the operating surgeon or identified by the trial physician screening postoperative patients on the neurosurgical ward. Patients were recruited as early postoperatively as possible, commonly on postoperative day two or three. Exclusion criteria were age below 18 years, pregnancy, occurrence of more than one generalized seizure per week, inability to provide written informed consent as well as contraindications for receiving MR-imaging or a TMS examination (for example sensitive metallic implants in direct vicinity to the stimulation site). Written informed consent was obtained before the first treatment session by a trial physician.

### Randomisation and masking

Patients were randomly assigned (1:1) to receive either sham-rTMS (control group) or verum- rTMS. In both groups, stimulation was followed by physiotherapy of the affected limbs according to the Bobath concept [12]. Randomisation was based on a computer-generated sequence without further stratification as created by the trial statistician. After the initial screening and enrollment of patients by the trial physician, patients were randomised by a study nurse. The study nurse was also responsible for performing the rTMS interventions throughout the trial but was otherwise not involved in assessment of the outcomes or patient communication. Blinding of the study nurse was not possible due to the chosen sham-rTMS approach. The trial physician, physiotherapists and patients were blinded to the group allocation of patients. All outcomes were assessed by blinded physicians or physiotherapists. Statistical analysis was performed following predefined analysis strategies without blinding to group allocation.

### Procedures

A T1-weighted structural MRI (TR 2300ms, TE 2.32ms, TI 900 ms, 9° flip angle, 256 × 256 matrix, 1 mm isotropic voxels, 192 slices, acquisition time: 5 min; Siemens Skyra 3T scanner, Siemens, Erlangen, Germany) acquired as part of the clinical routine was used as a subject-specific navigational dataset for the neuronavigated TMS (nTMS). NTMS was applied with a Nexstim NBS 5 stimulator (Nexstim, Helsinki, Finland) with a figure-of-eight coil (outer diameter 70mm). Muscle activity was recorded from the non-affected first dorsal interosseous muscle (FDI) via disposable Ag/AgCl surface electrodes (Neuroline 700; Ambu, Ballerup, Denmark) attached in a belly-tendon fashion. The ground electrode was placed on the left palmar wrist. The cortical representation of the FDI defined as point, electric field direction and angulation consistently eliciting the largest motor evoked potentials was recorded as intervention target. For this point, the resting motor threshold (RMT) was determined using the systems inbuilt automated threshold hunting algorithm [13]. During these baseline assessments, muscle activity was monitored to remain below 10µV.

Starting on the same day, patients received 7 sessions of week-daily rTMS (1Hz, 15 minutes, 900 pulses, 110% RMT) to the previously defined cortical target contralateral to the injury (Fig. 1) [8,10]. For sham-stimulation, a plastic adapter was placed onto the coil thus creating a 7cm space to the patient’s head. In this manner, the residual electric field reaching the cortex surface could be reduced to ≤ 5V/m. Patients were instructed to sit comfortably with relaxed muscles but were allowed to talk or perform subtle movements during stimulation. All patients received week-daily standardized physiotherapy of the affected limbs directly after rTMS. Daily rTMS sessions were aimed to take place at the same time each day.

**Fig. 1.**
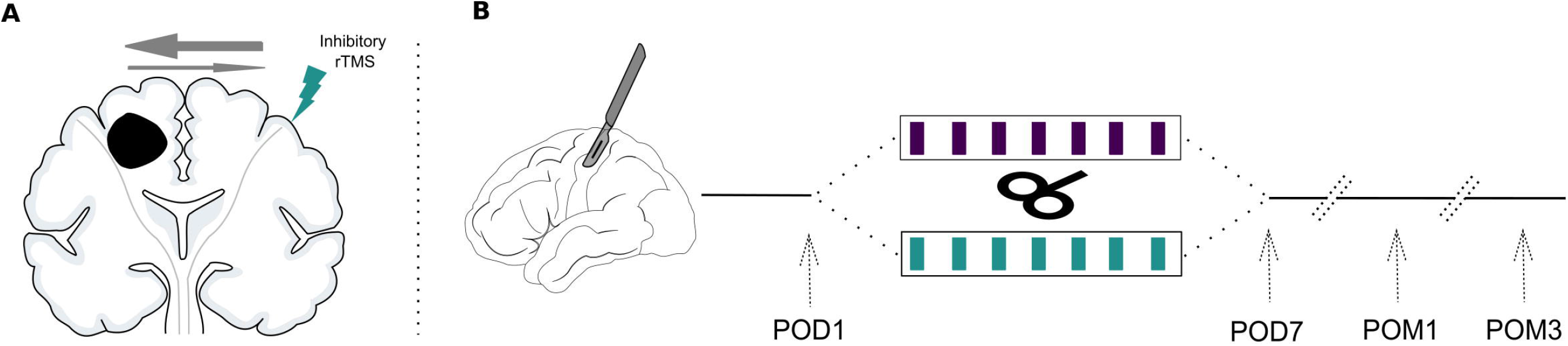
Study design. (A) Inhibitory rTMS is applied to the contralesional motor cortex to normalize the disturbed interhemispheric inhibitory balance due to the unihemispheric lesion. (B) Patients are recruited after resection of a supratentorial brain tumor and their baseline values are assessed (postoperative day 1; POD1). They are then randomized to receive either 7 sessions of week-daily rTMS or sham stimulation. Treatment effects are measured directly at the end of the interventional period on postoperative day 7 (POD7), one month postoperatively (POM1) and three months postoperatively (POM3).

### Outcomes

As primary outcome, the motor functioning domain of the Fugl Meyer Score (FMS; Part A-D, score between 0 [hemiplegic] and 66 [normal motor performance]) [14] for upper extremities was assessed by an experienced physiotherapist three months postoperatively. The FMS comprises a comprehensive assessment of muscle function including testing of reflexes, volitional movement with varying synergy levels and movement coordination. Outcomes were assessed before the first rTMS session (baseline), after the last rTMS session (POD7), after one month (POM1) and after three months (POM3). Fig. 1B visualizes the study timeline.

Secondary outcomes included: (i) FMS at POD7 and POM1. (ii) Ability to complete the Nine-Hole Peg Test (binary variable [yes, no]) [15]. Initially, time to complete the test was recorded. However, most patients were not able to perform the test due to a higher-grade paresis. Thus, we decided to binarize the Nine-Hole Peg Test score (able to complete test/not able to complete test). (iii) Number of finger taps achieved with the paretic index finger in 10 seconds. (iv) Overall functional impairment as measured with the Karnofsky Performance Status (KPS; score between 0 [dead] and 100 [healthy, no symptoms or signs of disease]) [16]. (v) Muscle strength of the distal and proximal paretic upper limb muscles measured with the BMRC (score between 0 [no muscle contraction] and 5 [normal muscle strength]) [17]. (vi) Overall score in the National Institute of Health Stroke Scale (NIHSS; score between 0 [no stroke symptoms] and 42 [severe stroke]) [18]. (vii) Health-related quality of life measured with the EORTC-QLQ-C30 (score between 0 [worst outcome] and 100 [best outcome]) [19]. This outcome was not assessed on postoperative day 1 but preoperatively. (viii) Duration between tumor resection and receiving adjuvant therapy (in weeks). If patients did not receive any adjuvant therapy until the last study follow-up three months postoperatively, a duration of 13 weeks was recorded. Outcomes (i) – (v) were assessed by a blinded physiotherapist, while outcomes (vi) – (viii) were quantified by a blinded physician.

Any event that was possibly, likely or highly likely related to the study intervention was recorded as study-related adverse event, while events that were unlikely to be related or not related to the study were classified as study-unrelated. Adverse events that led to a serious deterioration in patients’ health or death were defined as serious adverse events.

### Statistical analysis

Sample size calculation: Initially, a sample size of 15 patients per group was estimated to detect a difference in Fugl Meyer Scores between both groups (estimated effect size (Cohen’s d) of at least 0.74) with a power of 80% (two-sided significance level 0.05, independent samples t-test).

Data were analysed in an intention to treat (ITT) framework. Missing values for patients who died or were on palliative care were imputed by single imputation using worst possible values, since these missing values were informative missings. Similarly for patients who were not treated because of no deficits in motor function, missing values were imputed using single imputation by best possible values. For all other missing values, we used multiple imputation by chained equations (MICE) for estimation of missing values during follow up assuming missing at random (MAR) or missing completely at random (MCAR). Thirty complete datasets were used for the analyses of primary and secondary outcomes.

Differences between both treatment groups for each linear outcome were evaluated via linear mixed models (random intercept models to account for repeated measures within patients) adjusted for baseline values and including a time point by group interaction term to account for differential group differences at different time points. BMRC scores were analysed using mixed ordinal logistic regression models (random intercept models) adjusted for baseline values and including an interaction term of time point * group. Generalized estimation equation (GEE) binary logistic regression models adjusted for baseline performance and including time point and group allocation as well as the interaction term for group * time point were used to analyse completion of the Nine-Hole Peg Test (yes/no) and a binarized variable for finger tapping (0-20 / 21+). To test the effect of the intervention on the time until adjuvant treatment was started, times (weeks) of both groups were compared with a Wilcoxon rank sum test. The primary hypothesis was tested at a two-sided significance level α of 0.05. All secondary hypotheses were tested exploratory. Interpretation is based on effect estimates and 95%CI.

Additionally, we performed a subgroup analysis only in patients with a motor eloquent ischemia on the postoperative MRI to compare results to Ille et al. [9]. For the models for the finger tapping (in the total sample and the subgroup analysis) and distal BMRC scores (in the subgroup analysis) we omitted the interaction term of time*group since descriptive analyses did not reveal substantial differential treatment effects over time and since more complex models did not reach converge. These outcomes were therefore analysed only with a main effect for group and time point. As sensitivity analyses the same models as for the main analysis were repeated for the data before imputation (complete cases only) and after single imputation for informative missings.

Safety outcomes were analysed descriptively using summary statistics. Statistical analyses were performed in RStudio (Version 1.3.1073, http://www.rstudio.com/). The following R packages were used: base [20], mice [21], tidyverse [22], lme4 [23], ggeffects [24], ordinal [25], geepack [26].

## Results

Thirty patients were recruited between Feb 10, 2016 and Jul 22, 2021 and randomly assigned to the rTMS group and the control group. Nine patients in the rTMS group and 11 in the sham group completed the 3-month follow-up. Two patients died during the study period, two patients were on palliative care in a hospice. One patient had no therapy because the motor deficits resolved until the first therapy session. Details on the patient flow through different trial phases can be found in Fig. 2. All 30 patients were included in the analyses (Intention-to-treat) after single imputation for information missings and multiple imputation for MAR. Table 1 summarizes demographics and baseline characteristics of these patients.

**Fig. 2.**
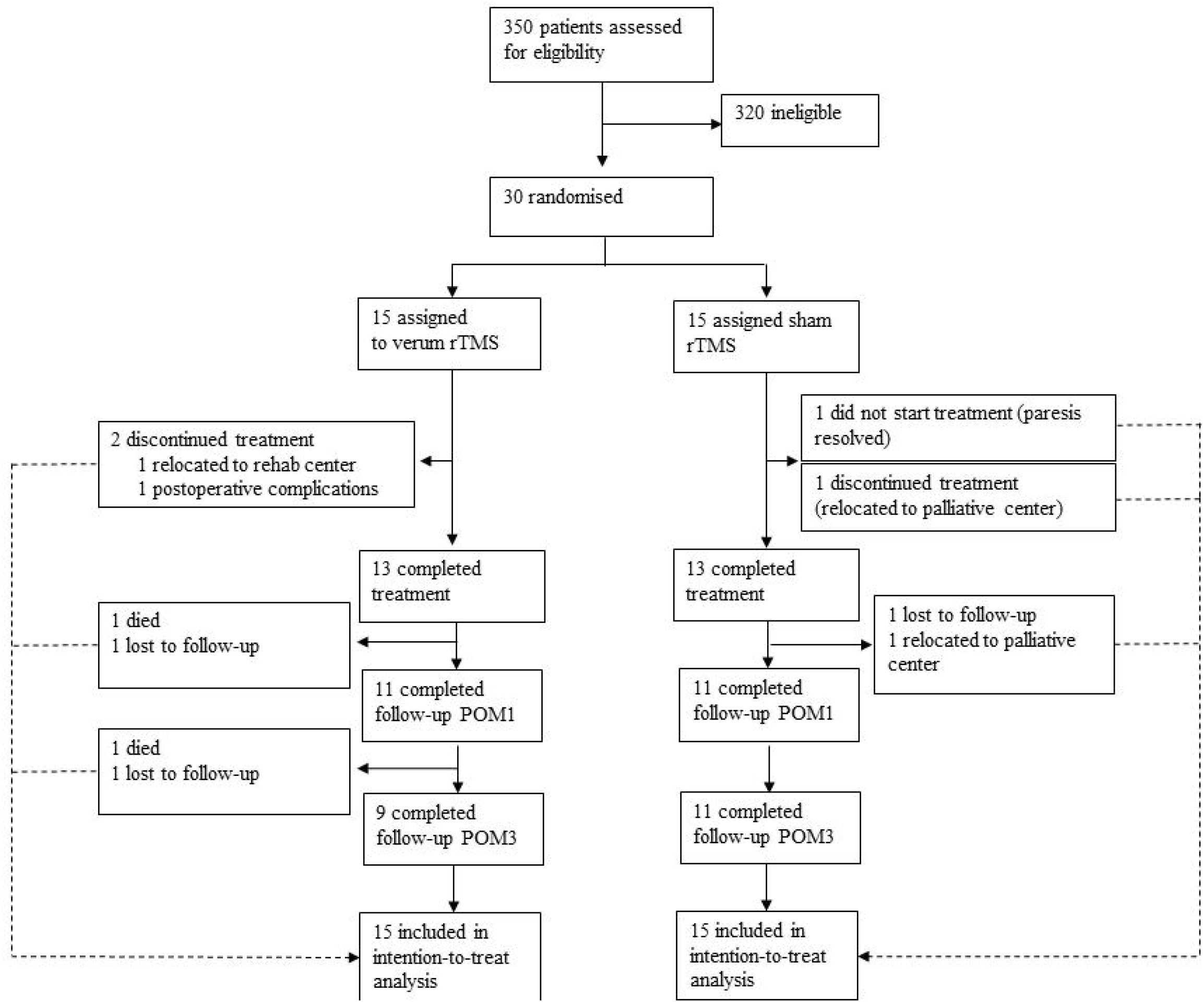
Patient flow throughout the trial. All patients were included in the intention-to-treat analysis and missing values were estimated using single imputations for informative missings and multiple imputations for missings at random (MAR) (see statistical analysis for details). POM1 = postoperative month 1, POM3 = postoperative month 3.

**Table 1.** Baseline characteristics by group. Data are assessed on POD1 and presented as mean (SD), n (%), or median (IQR). Deviations from 100% due to rounding. * One patient in rTMS and one in the sham group did not receive a postoperative MRI of sufficient quality to determine ischemia occurrence. ** One subject in the sham group did not perform the Finger Tapping test at baseline. *** Preoperative assessment

|  | rTMS group (n = 15) | Sham group (n = 15) | Overall (n = 30) |
| --- | --- | --- | --- |
| Age, years | 50 (11) | 54 (13) | 52 (12) |
| Sex |  |  |  |
| Female | 6 (40%) | 7 (47%) | 13 (43%) |
| Male | 9 (60%) | 8 (53%) | 17 (57%) |
| Time between surgery and first therapy, days | 3 (1) | 3 (2) | 3 (1) |
| Tumor Entity |  |  |  |
| GBM | 8 (53%) | 9 (60%) | 17 (57%) |
| Other glioma | 4 (27%) | 5 (33%) | 9 (30%) |
| Other | 3 (20%) | 1 (7%) | 4 (13%) |
| Tumor Location |  |  |  |
| Frontal | 6 (40%) | 3 (20%) | 9 (30%) |
| Parietal | 3 (20%) | 2 (13%) | 5 (17%) |
| Subcortical | 4 (27%) | 8 (53%) | 12 (40%) |
| Multilocular | 2 (13%) | 2 (13%) | 4 (13%) |
| Recurrence |  |  |  |
| De novo tumor | 10 (67%) | 11 (73%) | 21 (70%) |
| Recurrence | 5 (33%) | 4 (27%) | 9 (30%) |
| Subcortical ischemia* |  |  |  |
| None | 5 (33%) | 3 (21%) | 8 (29%) |
| Motor eloquent | 5 (33%) | 11 (79%) | 16 (57%) |
| Non motor eloquent | 4 (27%) | - | 4 (24%) |
| Fugl Meyer Score | 19 (1-41) | 3 (0-14) | 6 (0-22) |
| Nine-Hole Peg Test |  |  |  |
| Able to perform test | 5 (33%) | - | 5 (17%) |
| Unable to perform test | 10 (67%) | 15 (100%) | 25 (83%) |
| Number of finger Taps** | 0 (0-38) | 0 (0-0) | 0 (0-6) |
| Category ≤20 Taps | 10 (67%) | 14 | 24 (83%) |
| Category >20 Taps | 5 (33%) | - | 5 (17%) |
| Karnofsky Performance Index | 50 (40-75) | 40 (40-60) | 45 (40-60) |
| BMRC Score |  |  |  |
| Distal muscles | 1 (0-4) | 0 (0-1) | 0 (0-2) |
| Category 0 | 6 (40%) | 10 (67%) | 16 (53%) |
| Category 1-3 | 4 (27%) | 5 (33%) | 9 (30%) |
| Category 4-5 | 5 (33%) | - | 5 (17%) |
| Proximal muscles | 1 (0-4) | 0 (0-2) | 0 (0-2) |
| Category 0 | 7 (47%) | 10 (67%) | 17 (57%) |
| Category 1-3 | 4 (27%) | 5 (33%) | 9 (30%) |
| Category 4-5 | 4 (27%) | - | 4 (13%) |
| NIHSS Score | 7 (5-11) | 9 (6-11) | 9 (5-11) |
| Health-related quality of life (EORTC-QLQ-C30 Score)*** | 82 (67-84) | 82 (72-90) | 82 (69-87) |

### Postoperative motor outcome

All recorded outcomes for each timepoint are displayed in Fig. 3 on the single patient level together with the respective group averages.

**Fig. 3.**
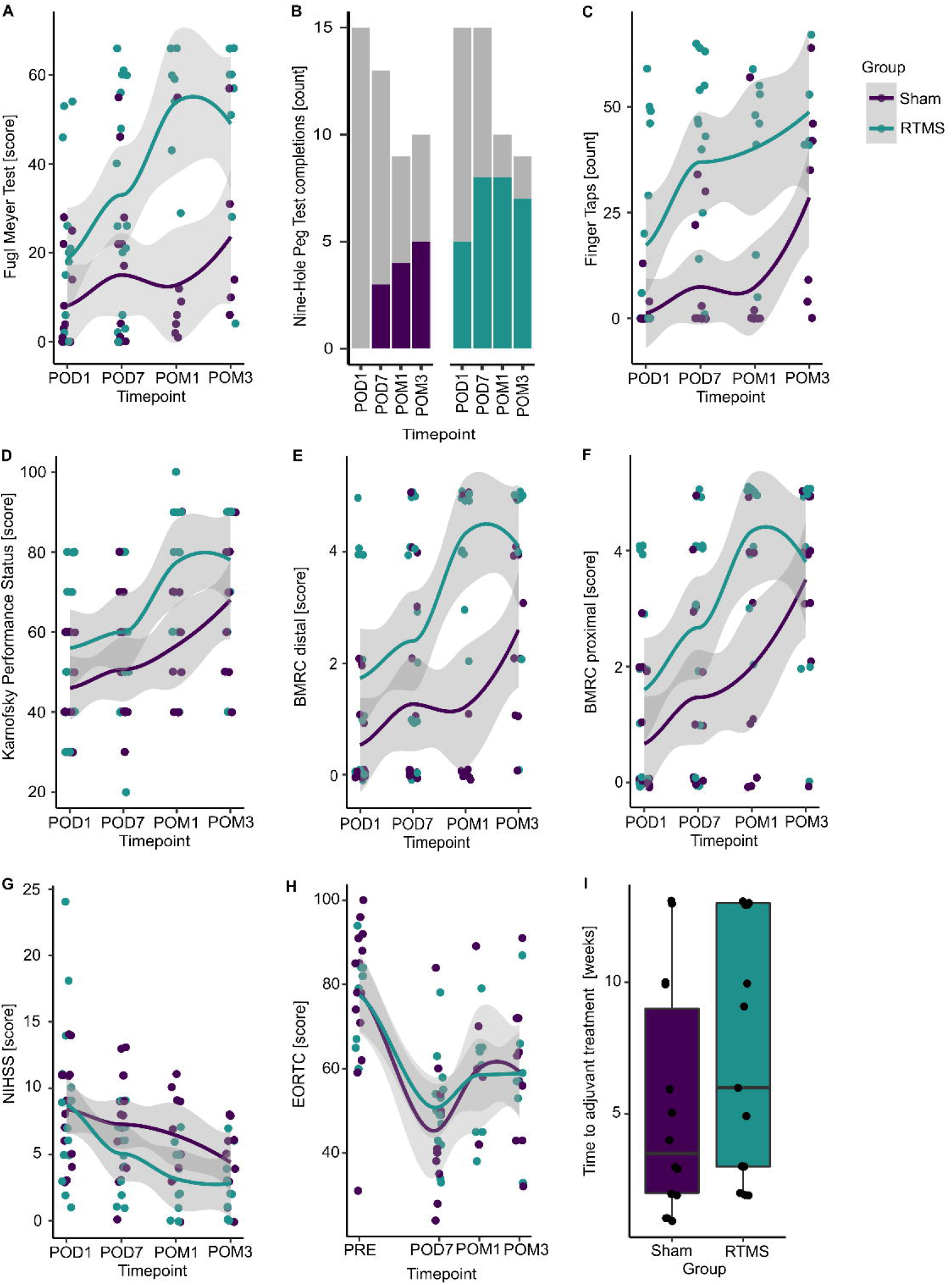
Treatment effects for all outcomes. Displayed are all recorded values (without imputations) for each timepoint separated by group (purple = sham, blue = rTMS). Colored points (A,C-I) represent single subject values. Colored lines (A, C-H) correspond to estimated group averages using locally estimated scatterplot smoothing (loess) with their respective 95% confidence intervals. (B) Colored bars display the number of patients being able to perform the task in each group and timepoint, while grey portions represent those subjects not being able complete it. (I) Boxplots visualize group differences in the time to adjuvant treatment. POD1 = postoperative day 1 (baseline), POD7 = postoperative day 7, POM1 = postoperative month 1, POM3 = postoperative month 3.

Three months postoperatively, patients in the rTMS group presented with slightly better Fugl Meyer Scores compared to the control group (5.1 [-16.0; 26.1]; p=0.631; Table 2). The model estimated mean Fugl Meyer Scores were 34.9 [20.2; 49.6] for the rTMS group versus 29.9 [15.1; 44.7] for the control group.

**Table 2.**
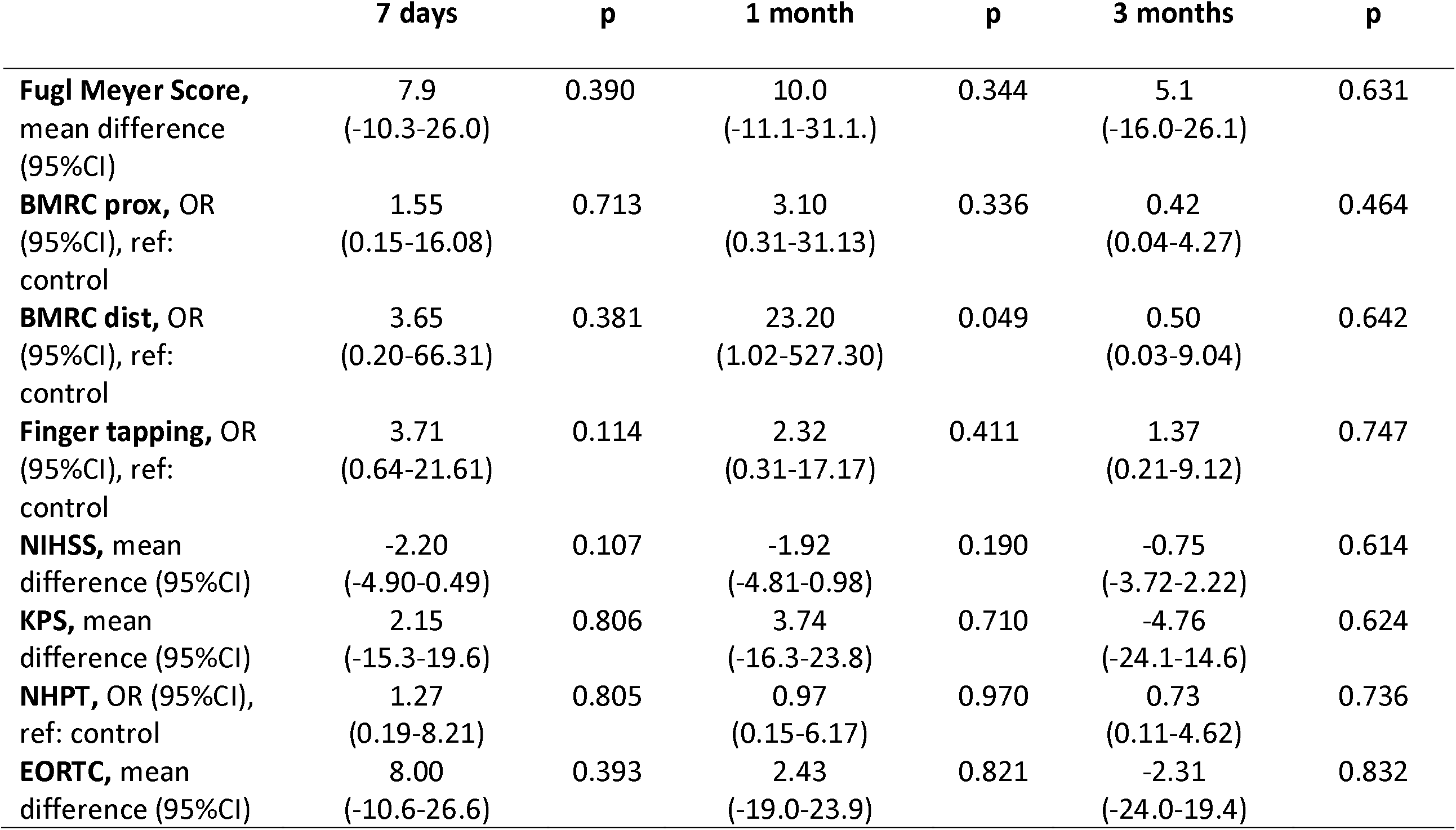
Postoperative outcomes. Treatment effects (mean group differences/odds ratios, 95% confidence intervals, p-values) for different outcomes (based on separate mixed models for each outcome, adjusted for baseline measures and including time point, group and interaction group*time point). Mixed models are calculated in multiple imputed datasets (m=30 data sets). N=30 individuals, 90 measures per model. prox = proximal muscles, dist = distal muscles, KPS = Karnofsky Performance Status, NHPT = Nine-Hole Peg Test.

|  | 7 days | p | 1 month | p | 3 months | p |
| --- | --- | --- | --- | --- | --- | --- |
| **Fugl Meyer Score,**<br>mean difference<br>(95%CI) | 7.9<br>(-10.3-26.0) | 0.390 | 10.0<br>(-11.1-31.1.) | 0.344 | 5.1<br>(-16.0-26.1) | 0.631 |
| **BMRC prox, OR**<br>(95%CI), ref:<br>control | 1.55<br>(0.15-16.08) | 0.713 | 3.10<br>(0.31-31.13) | 0.336 | 0.42<br>(0.04-4.27) | 0.464 |
| **BMRC dist, OR**<br>(95%CI), ref:<br>control | 3.65<br>(0.20-66.31) | 0.381 | 23.20<br>(1.02-527.30) | 0.049 | 0.50<br>(0.03-9.04) | 0.642 |
| **Finger tapping, OR**<br>(95%CI), ref:<br>control | 3.71<br>(0.64-21.61) | 0.114 | 2.32<br>(0.31-17.17) | 0.411 | 1.37<br>(0.21-9.12) | 0.747 |
| **NIHSS, mean**<br>difference (95%CI) | -2.20<br>(-4.90-0.49) | 0.107 | -1.92<br>(-4.81-0.98) | 0.190 | -0.75<br>(-3.72-2.22) | 0.614 |
| **KPS, mean**<br>difference (95%CI) | 2.15<br>(-15.3-19.6) | 0.806 | 3.74<br>(-16.3-23.8) | 0.710 | -4.76<br>(-24.1-14.6) | 0.624 |
| **NHPT, OR (95%CI),**<br>ref: control | 1.27<br>(0.19-8.21) | 0.805 | 0.97<br>(0.15-6.17) | 0.970 | 0.73<br>(0.11-4.62) | 0.736 |
| **EORTC, mean**<br>difference (95%CI) | 8.00<br>(-10.6-26.6) | 0.393 | 2.43<br>(-19.0-23.9) | 0.821 | -2.31<br>(-24.0-19.4) | 0.832 |

For the secondary outcomes, the BMRC score for distal upper extremity muscles was better at month 1 (OR: 23.20 [1.02; 527.30]; p=0.049) compared to the sham group. In contrast, only slight differences between the rTMS and sham group were observed for all remaining outcomes and timepoints (Table 2).

### Impact of motor eloquent ischemia

Looking only at patients with a motor eloquent ischemia on the postoperative MRI (n=16; 5 in rTMS group, 11 in sham group), the rTMS group presented with better NIHSS scores at postoperative day 7 (-3.95 [-7.68; 0.21]; p=0.039), month 1 (-4.38 [-8.12; 0.64]; p=0.024) and month 3 (-3.83 [-7.57; 0.097]; p=0.045). For all other outcomes and timepoints, only slightly better values were observed in the verum rTMS compared to the sham group (Supplementary table A.1).

### Sensitivity analysis

To quantify the impact of the multiple imputations on these results, we performed a sensitivity analysis on the treatment effects before imputation and after single imputation of informative missings only. The complete case analysis before imputations showed better outcomes in the rTMS group compared to the sham group for the BMRC for distal upper extremity muscles (OR: 511.00 [1.55-168746.16]; p=0.035). The analysis after single imputation showed no substantial differences between both groups. The complete results of the sensitivity analysis are summarized in supplementary table A.2.

### Adverse events

Generally, all patients tolerated the intervention well and completed stimulation at the designed intensity. The most common adverse events were headaches, followed my mild nausea and dizziness. These adverse events were judged to be related to the directly preceding surgery rather than the rTMS therapy and controlled with the standard postoperative patient management. Two serious adverse events were reported in two patients two weeks and three months postoperatively respectively, both leading to death of the patients. These serious adverse events were related to the underlying malignancy and growth of the tumor and judged unrelated to the intervention.

## Discussion

In this randomised, double-blind, sham-controlled, parallel-group study, we found that low-frequency rTMS of the unaffected primary motor cortex in tumor patients with postoperative motor deficits led to only slightly better Fugl Meyer scores three months postoperatively compared to sham stimulation. Analyses of secondary endpoints showed better hand motor function as measured with the BMRC in the rTMS group one month postoperatively compared to the control group. None of the other analyses of motor or neurological function or quality of life showed a substantial effect of the rTMS intervention. In the subgroup of patients with motor eloquent ischemia, rTMS additionally improved global neurological function at all timepoints. Results of the sensitivity analyses are in line with these findings. The intervention was generally well tolerated and no intervention-related serious adverse events occurred [27]. These findings suggest that low frequency rTMS can induce a facilitation of hand motor recovery specifically at early timepoints, while generalization of recovery to other muscle groups might require protocol adaptations.

The absence of substantial treatment effects at the long term follow up could be due to differences in timing of the adjuvant treatment and stays at rehabilitative centers both possibly impacting motor function. Further, highly malignant tumors may have early recurrences, causing secondary deterioration of motor function. It is also possible that the main effect of rTMS treatment is due to a faster compensation of the interhemispheric excitation level, which could explain the different treatment effects at different time points.

One other study [10] investigating the effect of the same intervention found significantly stronger improvements in Fugl Meyer, NIHSS and KPS scores after verum rTMS in brain tumor patients three months postoperatively. They did not assess early functional recovery, thus making comparison of recovery timelines between both studies difficult. They further limited their inclusion to tumor patients with subcortical ischemia within the corticospinal tract and preserved postoperative motor evoked potentials. Inclusion of patients only with preserved motor evoked potentials limits the study population to less severely affected patients, which might also explain higher effect sizes in this study compared to our results. However, the results of our subgroup analysis are in line with Ille et al. [10], suggesting patients with ischemia might be more susceptible to the treatment effects of rTMS.

This is supported by studies on rTMS therapy of motor deficits in stroke patients, providing level A evidence for the efficacy of low-frequency rTMS to the contralesional M1 for recovery of upper extremity motor function [8] in the postacute stage. Results for the chronic phase after stroke are mixed, thus suggesting the benefit of early interventions. These results support our approach to start the rTMS therapy at the earliest stage to facilitate recovery in the acute phase after injury.

Coordinated activation of both motor cortices is crucial for normal motor function [4]. In glioma patients, a preoperative imbalance of motor cortical excitability has been predictive of postoperative motor impairments [1]. Specifically, a lower excitability of the of the affected hemisphere compared to the unaffected one presented with a higher rate of postoperative motor deficits. Existing preoperative deficits in these patients did not recover postoperatively. These findings suggest that a disturbance of transcallosal activity might play a role in the development of motor deficits in tumor patients similar to earlier findings in stroke patients [28]. Thus, a normalization of interhemispheric excitability via application of rTMS might be beneficial for recovery of motor deficits also in tumor patients without ischemia. Increasing the target population would make the intervention clinically more relevant and profitable, although results from this study suggest a reduction in effect sizes in this group compared to other studies [10]. Future studies should examine these patient groups in more detail to delineate responder groups and potentially modify the treatment parameters in groups that are currently responding less good.

It could be criticized that assuming an increased activity of the unaffected motor cortex to be a maladaptive process might neglect functional reorganization and compensatory processes in slow-growing tumors. Recent neuromodulatory approaches focus on individualization of treatment based on detailed network analyses [29] and brain state dependent stimulation [30]. Implementing a patient-tailored treatment protocol was not possible in the present study due to limited availability of patients postoperatively and the acuteness of injury precluding, for example, stimulation of the affected hemisphere. Future studies should assess the impact of functional reorganization induced by tumor growth as well as potential differences in the degree of interhemispheric disbalance between patients on treatment success.

In theory, applying more stimulation sessions might increase efficacy of the intervention. Stimulation duration in this study was limited by capacities of the neurosurgical ward, subsequent adjuvant treatment or transfer to rehabilitation clinics. Future studies could investigate treatment protocols using accelerated rTMS, where multiple treatment sessions are applied per day [31], or the usefulness of booster sessions during the following postoperative weeks as they are used in patients with depression [8]. Similarly, the optimal target for stimulation should be further investigated to support the generalization of the induced recovery to other muscle groups.

Importantly, in neuromodulation studies adequate blinding of patients is challenging [32,33]. We used a sham control identical to verum rTMS, but with a plastic adapter between the TMS coil and patients’ heads. This approach reduces the electric field reaching the cortex surface, while providing the same sound and sensation of the coil on the head. In our experience, this blinding procedure is sufficient in patients naïve to rTMS as they are not informed whether they will feel muscle twitches during the intervention.

## Conclusions

Low-frequency rTMS might be a promising add-on treatment to standard physical therapy in patients after brain tumor resection. The treatment effect seems to be most prominent in stimulated muscle groups and more pronounced in patients with a motor eloquent postoperative ischemia. Yet more research is needed to increase effect sizes and identify patient groups with higher responsiveness to rTMS. Once this is achieved, a facilitation of motor recovery could reduce the disease burden in patients, leading to a better tolerability of adjuvant treatments, faster return to work and higher quality of life.

## Data Availability

Data produced in the present study cannot be shared due to data protection restrictions.

## CRediT author statement

Melina Engelhardt: Formal analysis, Investigation, Data Curation, Writing – Original Draft, Visualization, Project administration. Heike Schneider: Investigation, Project administration, Writing – Review & Editing. Jan Reuther: Conceptualization, Methodology, Writing – Review & Editing. Ulrike Grittner: Formal analysis, Writing – Review & Editing. Peter Vajkoczy: Conceptualization, Methodology, Writing – Review & Editing. Thomas Picht: Conceptualization, Methodology, Writing – Review & Editing, Supervision. Tizian Rosenstock: Conceptualization, Methodology, Investigation, Writing – Review & Editing, Supervision.

## Acknowledgements

Dr. Rosenstock is participant in the BIH Charité Digital Clinician Scientist Program funded by the Charité – Universitätsmedizin Berlin, and the Berlin Institute of Health at Charité (BIH).

## References

[1] Rosenstock T, Grittner U, Acker G, Schwarzer V, Kulchytska N, Vajkoczy P, et al. Risk stratification in motor area–related glioma surgery based on navigated transcranial magnetic stimulation data. J Neurosurg 2017;126:1227–37. https://doi.org/10.3171/2016.4.JNS152896.

[2] Zetterling M, Elf K, Semnic R, Latini F, Engström ER. Time course of neurological deficits after surgery for primary brain tumours. Acta Neurochir (Wien) 2020;162:3005–18. https://doi.org/10.1007/s00701-020-04425-3.

[3] Jakola AS, Gulati S, Weber C, Unsgård G, Solheim O. Postoperative deterioration in health related quality of life as predictor for survival in patients with glioblastoma: A prospective study. PLoS One 2011;6. https://doi.org/10.1371/journal.pone.0028592.

[4] Geffen GM, Jones DL, Geffen LB. Interhemispheric control of manual motor activity. Behavioural Brain Research 1994;64:131–40. https://doi.org/10.1016/0166-4328(94)90125-2.

[5] Murase N, Duque J, Mazzocchio R, Cohen LG. Influence of interhemispheric interactions on motor function in chronic stroke. Ann Neurol 2004;55:400–9. https://doi.org/10.1002/ana.10848.

[6] Duque J, Hummel F, Celnik P, Murase N, Mazzocchio R, Cohen LG. Transcallosal inhibition in chronic subcortical stroke. Neuroimage 2005;28:940–6. https://doi.org/10.1016/j.neuroimage.2005.06.033.

[7] Fitzgerald PB, Daskalakis ZJ. The Mechanism of Action of rTMS. Repetitive Transcranial Magnetic Stimulation Treatment for Depressive Disorders, Springer Berlin Heidelberg; 2013, p. 13–27. https://doi.org/10.1007/978-3-642-36467-9_3.

[8] Lefaucheur JP, Aleman A, Baeken C, Benninger DH, Brunelin J, di Lazzaro V, et al. Evidence-based guidelines on the therapeutic use of repetitive transcranial magnetic stimulation (rTMS): An update (2014–2018). Clinical Neurophysiology 2020;131:474–528. https://doi.org/10.1016/j.clinph.2019.11.002.

[9] Piil K, Juhler M, Jakobsen J, Jarden M. Controlled rehabilitative and supportive care intervention trials in patients with high-grade gliomas and their caregivers: A systematic review. BMJ Support Palliat Care 2016;6:27–34. https://doi.org/10.1136/bmjspcare-2013-000593.

[10] Ille S, Kelm A, Schroeder A, Albers LE, Negwer C, Butenschoen VM, et al. Navigated repetitive transcranial magnetic stimulation improves the outcome of postsurgical paresis in glioma patients – A randomized, double-blinded trial. Brain Stimul 2021;14:780–7. https://doi.org/10.1016/j.brs.2021.04.026.

[11] World Medical Association. World Medical Association Declaration of Helsinki: Ethical Principles for Medical Research Involving Human Subjects. JAMA 2013;310:2191. https://doi.org/10.1001/jama.2013.281053.

[12] Bobath B. Treatment principles and planning in cerebral palsy. Physiotherapy 1963;49:122–4.

[13] Engelhardt M, Schneider H, Gast T, Picht T. Estimation of the resting motor threshold (RMT) in transcranial magnetic stimulation using relative-frequency and threshold-hunting methods in brain tumor patients. Acta Neurochir (Wien) 2019;161:1845–51. https://doi.org/10.1007/s00701-019-03997-z.

[14] Fugl-Meyer AR, Jääskö L, Leyman I, Olsson S, Steglind S. The post-stroke hemiplegic patient. a method for evaluation of physical performance. Scand J Rehabil Med 1975;7:13–31.

[15] Kellor M, Frost J, Silberberg N, Iversen I, Cummings R. Hand strength and dexterity. Am J Occup Ther 1971;25:77–83.

[16] Schag CC, Heinrich RL, Ganz PA. Karnofsky performance status revisited: reliability, validity, and guidelines. Journal of Clinical Oncology 1984;2:187–93. https://doi.org/10.1200/JCO.1984.2.3.187.

[17] Compston A. Aids to the Investigation of Peripheral Nerve Injuries. Medical Research Council: Nerve Injuries Research Committee. His Majesty’s Stationery Office: 1942; pp. 48 (iii) and 74 figures and 7 diagrams; with Aids to the Examination of the Peripheral Nervous System. By Michael O’Brien for the Guarantors of Brain. Saunders Elsevier: 2010; pp. [8] 64 and 94 Figures. Brain 2010;133:2838–44. https://doi.org/10.1093/brain/awq270.

[18] Brott T, Adams HP, Olinger CP, Marler JR, Barsan WG, Biller J, et al. Measurements of acute cerebral infarction: a clinical examination scale. Stroke 1989;20:864–70. https://doi.org/10.1161/01.STR.20.7.864.

[19] Aaronson NK, Ahmedzai S, Bergman B, Bullinger M, Cull A, Duez NJ, et al. The European Organization for Research and Treatment of Cancer QLQ-C30: A Quality-of-Life Instrument for Use in International Clinical Trials in Oncology. JNCI Journal of the National Cancer Institute 1993;85:365–76. https://doi.org/10.1093/jnci/85.5.365.

[20] R Core Team. R: A language and environment for statistical computing 2021.

[21] Buuren S van, Groothuis-Oudshoorn K. mice: Multivariate Imputation by Chained Equations in R. J Stat Softw 2011;45. https://doi.org/10.18637/jss.v045.i03.

[22] Wickham H, Averick M, Bryan J, Chang W, McGowan L, François R, et al. Welcome to the Tidyverse. J Open Source Softw 2019;4:1686. https://doi.org/10.21105/joss.01686.

[23] Bates D, Mächler M, Bolker B, Walker S. Fitting Linear Mixed-Effects Models Using lme4. J Stat Softw 2015;67. https://doi.org/10.18637/jss.v067.i01.

[24] Lüdecke D. ggeffects: Tidy Data Frames of Marginal Effects from Regression Models. J Open Source Softw 2018;3:772. https://doi.org/10.21105/joss.00772.

[25] Christensen RHB. ordinal - Regression Models for Ordinal Data. R package version 2022.11-16 2022.

[26] Halekoh U, Højsgaard S, Yan J. The R Package geepack for Generalized Estimating Equations. J Stat Softw 2006;15. https://doi.org/10.18637/jss.v015.i02.

[27] Rossi S, Antal A, Bestmann S, Bikson M, Brewer C, Brockmöller J, et al. Safety and recommendations for TMS use in healthy subjects and patient populations, with updates on training, ethical and regulatory issues: Expert Guidelines. Clinical Neurophysiology 2021;132:269–306. https://doi.org/10.1016/j.clinph.2020.10.003.

[28] Hsu W-Y, Cheng C-H, Liao K-K, Lee I-H, Lin Y-Y. Effects of Repetitive Transcranial Magnetic Stimulation on Motor Functions in Patients With Stroke. Stroke 2012;43:1849–57. https://doi.org/10.1161/STROKEAHA.111.649756.

[29] Menardi A, Momi D, Vallesi A, Barabási A-L, Towlson EK, Santarnecchi E. Maximizing brain networks engagement via individualized connectome-wide target search. Brain Stimul 2022;15:1418–31. https://doi.org/10.1016/j.brs.2022.09.011.

[30] Zrenner C, Desideri D, Belardinelli P, Ziemann U. Real-time EEG-defined excitability states determine efficacy of TMS-induced plasticity in human motor cortex. Brain Stimul 2018;11:374–89. https://doi.org/10.1016/j.brs.2017.11.016.

[31] Engelhardt M, Kimmel J, Raffa G, Conti A, Picht T. Safety and Tolerability of Accelerated Low-Frequency Repetitive Transcranial Magnetic Stimulation Over the Primary Motor Cortex–A Pilot Study. Front Neurosci 2022;16. https://doi.org/10.3389/fnins.2022.793742.

[32] Grasin E, Loginov I, Masliukova A, Smirnov N. Realistic sham TMS. Brain Stimul 2019;12:418. https://doi.org/10.1016/j.brs.2018.12.353.

[33] Duecker F, Sack AT. Rethinking the role of sham TMS. Front Psychol 2015;6. https://doi.org/10.3389/fpsyg.2015.00210.

