## Supplementary Material for "Low-frequency repetitive transcranial magnetic stimulation in patients with motor deficits after brain tumor resection: a randomised, double-blind, sham-controlled trial"

| | group difference for models without interaction of group*time | p | 7 days | p | 1 month | p | 3 months | p |
|---|---|---|---|---|---|---|---|---|
| **Fugl Meyer Score,** mean difference (95%CI) | -- | | 13.78 (-9.50-37.06) | 0.351 | 18.34 (-11.40-48.04) | 0.215 | 9.47 (-19.40-38.30) | 0.505 |
| **BMRC prox,** OR (95%CI), ref: control | -- | | 4.55 (0.19-109.8) | 0.351 | 2.20 (0.10-47.50) | 0.615 | 0.48 (0.02-11.10) | 0.646 |
| **BMRC dist,** OR (95%CI), ref: control | 13.6 (0.38-484) | 0.152 | -- | | -- | | -- | |
| **Finger tapping,** OR (95%CI), ref: control | 2.69 (0.24-30.8) | 0.420 | -- | | -- | | -- | |
| **NIHSS,** mean difference (95%CI) | -- | | -3.95 (-7.68-0.21) | 0.039 | -4.38 (-8.12-0.64) | 0.024 | -3.83 (-7.57-0.097) | 0.045 |
| **KPS,** mean difference (95%CI) | -- | | 8.34 (-16.2-32.9) | 0.488 | 17.97 (-11.8-47.7) | 0.223 | 11.84 (-17.7-41.4) | 0.415 |
| **NHPT,** OR (95%CI), ref: control | -- | | 1.23 (-1.88-4.34) | 0.437 | 0.55 (-2.48-3.58) | 0.720 | 0.30 (-2.59-3.19) | 0.839 |
| **EORTC,** mean difference (95%CI) | -- | | 13.0 (-15.1-41.1) | 0.354 | 12.7 (-15.8-41.1) | 0.373 | 14.9 (-15.0-44.8) | 0.320 |

**Table A.1 Impact of motor eloquent ischemia.** Treatment effects (mean group differences/odds ratios, 95% confidence intervals, p-values) for different outcomes (based on separate models for each outcome, adjusted for baseline measures and including time point, group and interaction group*time point) in the subgroup of patients with motor eloquent ischemia (n=16; 5 in rTMS group, 11 in sham group; 48 measures per model). Models for the finger tapping and distal BMRC scores did not converge when the interaction term for group * time point was included. They were therefore analysed only with a main effect for group and time point. Estimates are based on mixed models or GEEs. prox = proximal muscles, dist = distal muscles, KPS = Karnofsky Performance Status, NHPT = Nine-Hole Peg Test.

**Before imputation (complete case analysis)**

| | N individuals | N measures | group difference for models without interaction of group*time | p | 7 days | p | 1 month | p | 3 months | p |
|---|---|---|---|---|---|---|---|---|---|---|
| **Fugl Meyer Score,** mean difference (95%CI) | 27 | 52 | -- | | -5.78 (-19.00-7.43) | 0.378 | -15.22 (-31.83-1.40) | 0.072 | 2.33 (-15.00-19.66) | 0.788 |
| **KG prox,** OR (95%CI), ref: control | 29 | 71 | -- | | 1.65 (0.18-15.23) | 0.658 | 3.19 (0.23-45.53) | 0.384 | 0.15 (0.01-2.35) | 0.175 |
| **KG dist,** OR (95%CI), ref: control | 30 | 71 | -- | | 8.62 (0.07-1078.83) | 0.382 | 511.00 (1.55-168746.16) | 0.035 | 0.42 (0.002-80.65) | 0.747 |
| **Finger tapping,** OR (95%CI), ref: control | 27 | 54 | 6.29 (0.94-42.00) | 0.058 | -- | | **--** | | **--** | |
| **NIHSS,** mean difference (95%CI) | 29 | 70 | -- | | -2.09 (-4.21-0.02) | 0.053 | -1.91 (-4.21-0.39) | 0.101 | -0.57 (-2.89-1.74) | 0.621 |
| **KPS,** mean difference (95%CI) | 30 | 71 | **--** | | 2.58 (-8.26-13.41) | 0.633 | 9.15 (-2.81-21.10) | 0.131 | -0.46 (-12.55-11.64) | 0.902 |
| **NHPT,** OR (95%CI), ref: control | 30 | 66 | -- | | 1.43 (0.22-9.26) | 0.708 | 1.88 (0.20-17.27) | 0.579 | 1.00 (0.10-10.17) | 1.000 |
| **EORTC QoL,** mean difference (95%CI) | 21 | 47 | **--** | | 2.19 (-11.60-15.92) | 0.748 | 4.42 (-12.70-21.58) | 0.605 | 3.10 (-13.50-19.67) | 0.707 |

| | | | | | | | | | | |
|---|---|---|---|---|---|---|---|---|---|---|
| **After single imputation of informative missings** | | | | | | | | | | |
| **Fugl Meyer Score,** mean difference (95%CI) | 27 | 59 | -- | | 5.50 (-9.37-20.37) | 0.457 | 7.44 (-10.12-25.00) | 0.398 | -4.23 (-22.09-13.60) | 0.636 |
| **BMRC prox,** OR (95%CI), ref: control | 30 | 79 | -- | | 1.51 (0.12-19.20) | 0.752 | 2.65 (0.18-39.70) | 0.481 | 0.21 (0.01-3.20) | 0.258 |
| **BMRC dist,** OR (95%CI), ref: control | 30 | 79 | -- | | 4.90 (0.10-234.00) | 0.424 | 46.90 (0.55-4030.00) | 0.090 | 0.20 (0.01-12.00) | 0.464 |
| **Finger tapping,** OR (95%CI), ref: control | 28 | 61 | 3.86 (0.59-25.10) | 0.158 | -- | | -- | | -- | |
| **NIHSS,** mean difference (95%CI) | 29 | 78 | -- | | -2.08 (-4.68-0.52) | 0.114 | -1.73 (-4.44-0.99) | 0.207 | -7.13 (-24.71-10.44) | 0.419 |
| **KPS,** mean difference (95%CI) | 30 | 79 | -- | | 1.12 (-15.30-17.53) | 0.891 | 3.84 (-13.88-21.55) | 0.666 | -0.46 (-12.55-11.64) | 0.940 |
| **NHPT,** OR (95%CI), ref: control | 30 | 75 | -- | | 1.43 (0.22-9.26) | 0.708 | 0.90 (0.13-6.08) | 0.914 | 0.58 (0.08-4.39) | 0.601 |
| **EORTC,** mean difference (95%CI) | 23 | 56 | -- | | 6.41 (-11.60-24.44) | 0.476 | 4.89 (-15.60-25.41) | 0.633 | -4.69 (-24.00-14.59) | 0.626 |

**Table A.2 Sensitivity analyses.** Treatment effects (mean group differences/odds ratios, 95% confidence intervals, p-values) for different outcomes before imputation and after single imputation for informative missings (based on separate models for each outcome, adjusted for baseline measures and including time point, group and interaction group*time point). Models for the finger tapping did not converge when the interaction term for group * time point was included. They were therefore analysed only with a main effect for group and timepoint. Estimates are based on mixed models or GEEs. prox = proximal muscles, dist = distal muscles, KPS = Karnofsky Performance Status, NHPT = Nine-Hole Peg Test.
